# Improvement of language function after C7 neurotomy at the intervertebral foramen in patients with chronic post-stroke aphasia: a phase I cohort study

**DOI:** 10.1101/2023.03.22.23287523

**Authors:** Juntao Feng, Xingyi Ma, Ruiping Hu, Minzhi Lv, Tie Li, Peiyang Li, Wenjun Qi, Miaomiao Xu, Jingrui Yang, Yundong Shen, Wendong Xu

## Abstract

**Background:** Post-stroke aphasia is a common but intractable sequela which still needs new and more effective treatments. Evidence from follow-ups after contralateral seventh cervical nerve transfer surgery indicated that nerve transection leads to immediate language improvements in patients with right post-stroke aphasia.

**Objective:** Through a prospective cohort design, this study aims to prove that C7 neurotomy at the intervertebral foramen (NC7) combined with a 3-week intensive speech and language therapy (iSLT) can improve the language function in post-stroke aphasia patients.

**Methods:** In this study, patients aged over 18 years old and had been diagnosed with post-stroke aphasia for 1 year or longer were included. Primary outcomes were the change in the ability to retrieve personally relevant words in Boston Naming Test (BNT) with follow-up assessment after three-weeks’ iSLT post-operatively. As well as several secondary outcome measures including the Western Aphasia Battery (WAB), daily communication abilities (measured by the Communication Activities of Daily Living Third Edition [CADL-3]) and Fugl-Meyer of upper limb part (UEFM).

**Results:** The average increase of BNT score was 11.2 points from baseline to 3 weeks post-operatively (P=0.001, 95%CI: 8.1-14.1). The WAB and CADL-3 assessment showed 9.4, 10.4 points increasing in average (P<0.005, 95%CI: 4.6 to 14.1; P<0.001, 95%CI:6.7 to 14.1) from baseline to 4-week follow-up, respectively. The mean difference from baseline to 3 weeks post-operatively in UEFM score decreased 0.8 points (95% CI: -3.2 to 1.6; p<0.405).

**Conclusions:** NC7 plus iSLT significantly improved the language function in patients with post-stroke aphasia, and did not significantly affect the motor function of the right limb. The mechanism of this surgery needs to be further explored.

## INTRODUCTION

Aphasia refers to a collection of acquired receptive and expressive language deficits [1]. It is a common sequela of stroke, frequently occurring in patients with left-sided hemiparesis [2, 3]. As a devastating condition to most stroke patients, aphasia affects nearly every social activity and interaction [4]. Current treatments focus on rehabilitation are reports of some improvement in language function, such as intensive speech and language therapy (iSLT) [5, 6], as well as low-frequency electrical stimulation therapy, repetitive transcranial magnetic stimulation (rTMS) [7], and transcranial direct current stimulation (tDCS) [8]. However, patients who reach the plateau stage have difficulty in further improving their language function [9].

We previously proposed that contralateral C7 transfer from the nonparalyzed side to the paralyzed side (contralateral C7 to C7 cross nerve transfer [CC7]) can effectively improve upper limb function [10, 11]. In the clinical follow-up of this surgery, post-stroke patients with complicated chronic aphasia surprisingly showed improvement in self-reported language function, not only in terms of fluency but also in terms of increased naming ability. This phenomenon was firstly found by patients’ self-reports and then confirmed through cohort follow-up study (unpublished yet). Since the language function improvement occurs within a short period of time after surgery (mostly within half a day after surgery) and the nerve regeneration has not yet begun, we think this improvement is thought to be closely related to neurotomy. There is also previous literature on improved language function in patients after selective posterior rhizotomies (SPR) surgery reported [12, 13]. These evidences indicated that that the improvement in language function is more likely related to the severed cervical seventh nerve on the affected side. Therefore, we propose a modified procedure for the treatment of chronic post-stroke aphasia, which is severing only the affected (right) cervical seventh nerve (the same as the procedure on the paralyzed side in CC7) at the intervertebral foramen (NC7), and observe whether there is an improvement in language function.

To verify the safety and preliminary effectiveness of NC7 at the intervertebral foramen on language impairment in patients with chronic post-stroke aphasia, we designed a prospective clinical small cohort study with BNT, WAB, Fugl-Meyer and other scales for language and motor function assessment.

## METHODS

### Study Design and Participants

This is a single center, prospective, explorative cohort study approved by the institutional review board (Institutional Review Board of Jing’an District Central Hospital Affiliated to Hua Shan hospital of Fudan University). Consents of the study were acquired for all patients participated in this study. In this study, we included five patients with post-stroke aphasia (> 1 year) who were planned to receive contralateral seventh cervical nerve neurotomy to improve the language function. The post-stroke aphasia is diagnosed by the Aphasia Quotient (< 93.8) calculated by the Western Aphasia Battery (WAB). The participants should be aged over 18 years old, right-handed, native Chinese speakers with adequate visual and auditory abilities to complete the assessments. Participants were excluded, if they had a speech and language disorder caused by other deficits or before the last disabling stroke (such as neurodegenerative diseases), if they were unable or unwilling to participant in the study, or if they were currently receiving intensive speech and language therapy within four weeks before screening.

### Clinical Assessments

In this article, we presented the language and upper extremity function of the patients at baseline (T0) and T1 follow-up which were evaluated before-intervention, after the surgery and 3-week iSLT plan, respectively. The primary outcome is the change in total score of the Boston naming test (BNT) scale from baseline to 3 weeks after the surgery. WAB, Communication Activities of Daily Third Edition (CADL-3), Fugl-Meyer of upper limb part (UEFM) and Modified Ashworth Score (MAS) were used as secondary outcomes.

For the language function, BNT is a classic measurement tool for evaluating language function, which shows a high concurrent validity with other standard naming ability assessment tools, and it is particularly suitable in the post-acute/chronic phase after stroke aphasia. In this study, we chose the validated Chinese version of BNT [14, 15]. The WAB applied in this study is consisted of four subtests, spontaneous speech, auditory comprehension, naming and repetition [16], and CADL-3 scale contains 50 items assessing functional communication skills in seven areas of adults with neurogenic communication disorders [17]. The types of aphasia of these patients were classified according to WAB-Aphasia quotation calculated by the scores of the WAB scale.

For the motor function, we used the UEFM to evaluate the paralyzed upper limb function, of which higher score indicated better motor function of the upper limb [18]. The spasticity of elbow and finger on the paralyzed side were also evaluated with MAS, with higher values indicating more spasticity [19]. Safety outcomes include any incidence related to the surgery or irrelevant but occurred after the surgery during the study period, such as bleeding, Infection, pain, numbness and so on. Patients will receive study information containing explicit details on whom to contact in case of an adverse event. Investigators will record all descriptions of adverse events during each follow-up visit.

Structural Magnetic Resonance Image(MRI)evaluation was applied in all 5 patients to detect the lesion site of left hemisphere [20]. Functional magnetic resonance image (fMRI) was also acquired but not analysed in this article.

### Interventions

#### NC7 at the intervertebral foramen

Make a 6-cm long longitudinal incision along the medial border of the sternocleidomastoid muscle on the right side after the cervical plexus under anaesthesia/general anaesthesia (depending on the patient’s preference and the anaesthesiologist’s risk assessment). Carefully separate the structure layer by layer and identify the brachial plexus by marking the C7 nerve with a vessel loop. Next, the C7 root is mobilized and sectioned as proximally (at the intervertebral foramen) as possible. Afterwards, the distal cut-end of the right C7 root was sutured to subcutaneous soft-tissues.

#### iSLT Rehabilitation

A 3-week iSLT was performed one-week after the surgery. Speech and language therapy with a therapist for at least 45 minutes, twice a day, 5 days per week. Additional 1 hour/day self-administrated language-specific training. Patients receive rehabilitation treatment in Shanghai Jing’an District Central Hospital, where qualified rehabilitation therapists perform the therapy.

### Statistics

For demographic data, function data and safety indicators at baseline and post-operative follow-up (week-4), descriptive statistics was performed. Categorical data are presented as frequency and percentage, and continuous data are described by standard deviation. A two-sample paired t-test was performed to compare within-group differences in BNT, WAB, CADL-3 and UEFM score, with 95% confidence intervals reported. P value is studied and P < 0.05 was considered statistically significant. The ratio of change in each scale were calculated as the mean change divided by the total score of the full scale or each subtest. SPSS 22.0 software was used for calculation.

## RESULTS

The 5 patients’ Demographic characteristics and baseline data was listed in Table 1. The lesion area of one slice from the MRI scanning was shown in Figure 1. In language function tests, we observed the BNT score showed significant increase from baseline to 4-week (mean change: 11.2, 95% CI: 8.1 to 14.3, P<0.001. The WAB and CADL-3 score both acquired significant increase from baseline to 4-week follow-up (mean change: 9.4, 95% CI: 4.6 to 14.1, P<0.005; mean change: 10.4, 95% CI: 6.7 to 14.1, P<0.001). In the motor function tests, the UEFM score showed decrease at 4-weeks follow-up (mean change: 0.8, 95%CI: -3.2 to 1.6, P< 0.405), but the difference did not meet the lowest significant level (P<0.05). The spasticity of the elbow, wrist and fingers decreased at 3 weeks after the surgery (see Table 2 and 3). Table 4 displayed the safety outcome and the adverse reactions that arose in each patient during this study. All patients’ tactile sensory threshold increased in thumb, index and middle fingers combined with numbness, and partial patients demonstrated pain, decreasing in muscle strength of the paralyzed side.

**Table 1.**
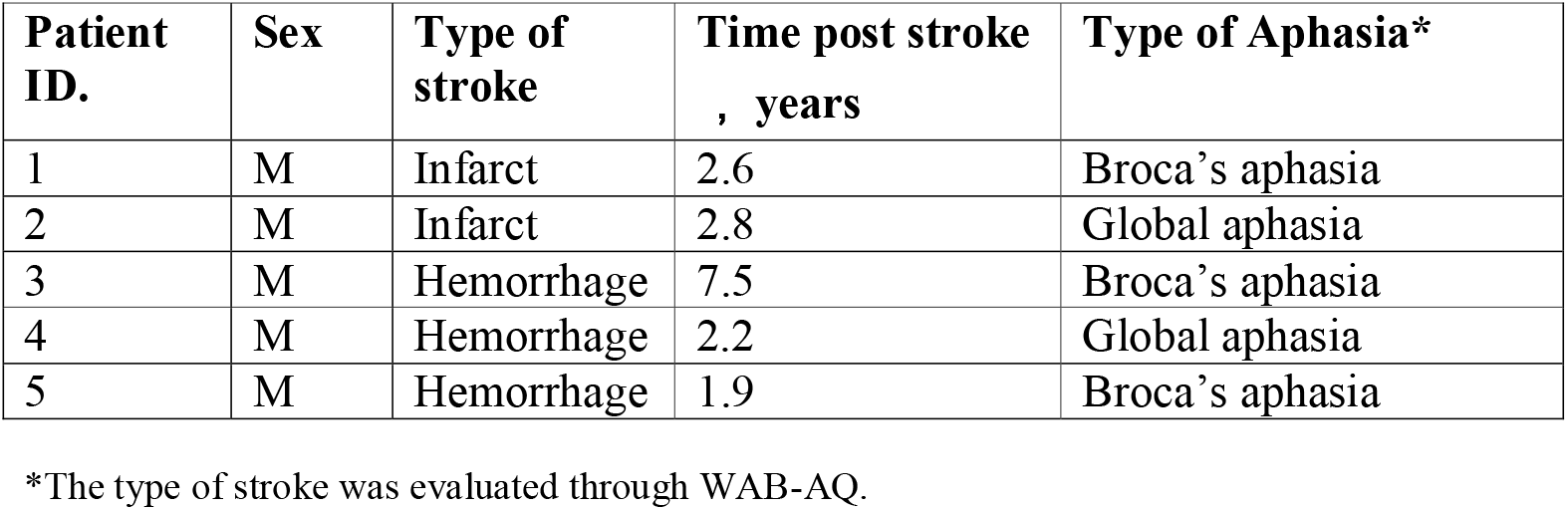
Demographic characteristics and baseline data of the patients included.

| **Patient ID.** | **Sex** | **Type of stroke** | **Time post stroke , years** | **Type of Aphasia\*** |
| --- | --- | --- | --- | --- |
| 1 | M | Infarct | 2.6 | Broca's aphasia |
| 2 | M | Infarct | 2.8 | Global aphasia |
| 3 | M | Hemorrhage | 7.5 | Broca's aphasia |
| 4 | M | Hemorrhage | 2.2 | Global aphasia |
| 5 | M | Hemorrhage | 1.9 | Broca's aphasia |

\*The type of stroke was evaluated through WAB-AQ.

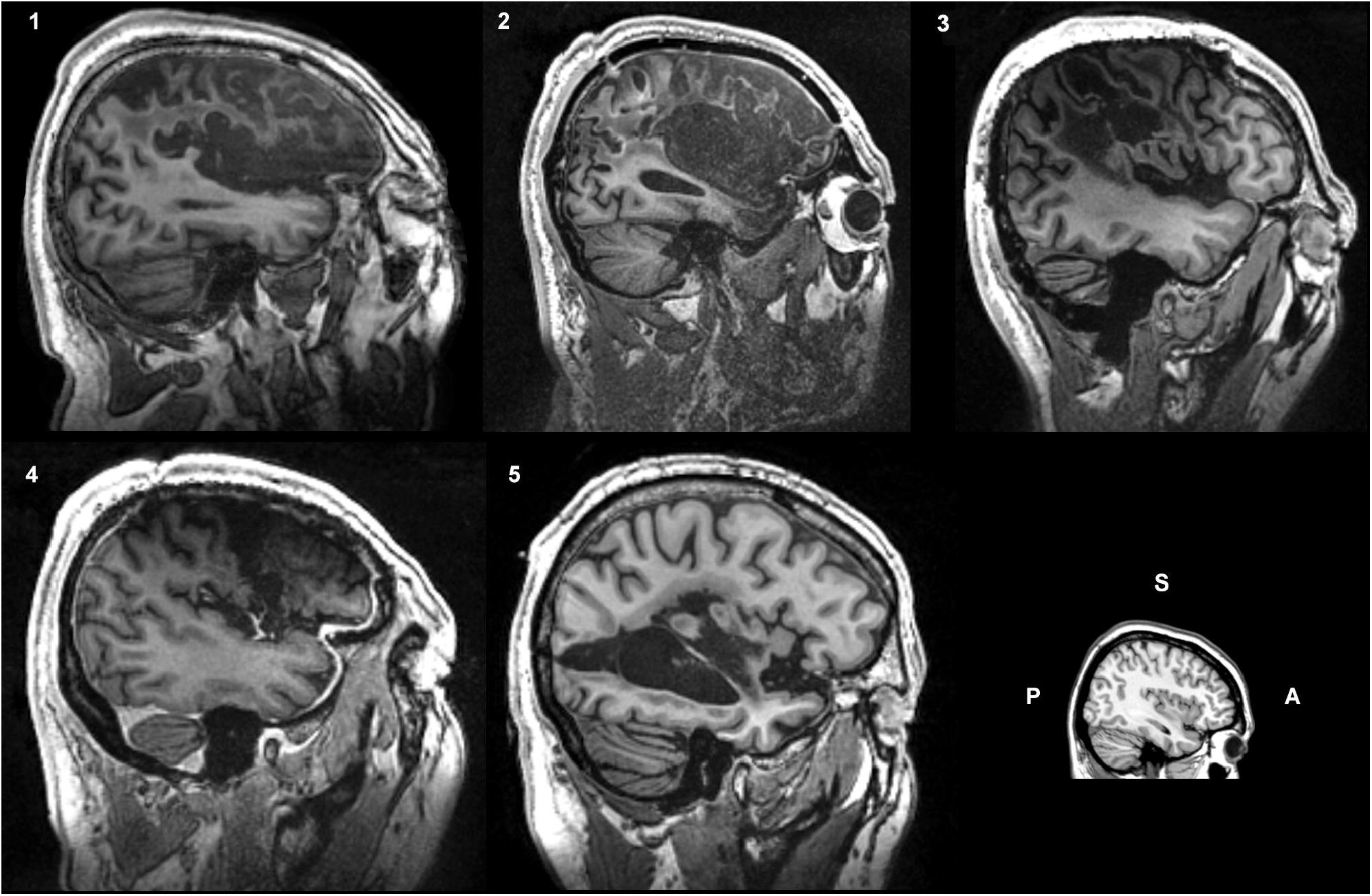

**Table 2.** Changes of the language and motor function evaluations from baseline to each follow-up.

| Patient ID |  | 1 | 2 | 3 | 4 | 5 | Average<br>(Mean±SD) |
| --- | --- | --- | --- | --- | --- | --- | --- |
| Outcomes |  |  |  |  |  |  |  |
| BNT-60 | Baseline | 13 | 4 | 39 | 1 | 6 | 12.6±15.4 |
|  | Week-3 | 25 | 13 | 54 | 10 | 17 | 23.8±17.8 |
|  | Change | 12 | 9 | 15 | 9 | 11 | 11.2±2.5 |
| WAB-AQ | Baseline | 41.8 | 27.2 | 57.8 | 11.0 | 52.0 | 38.0±19.0 |
|  | Week-3 | 51.2 | 31.8 | 69.8 | 17.8 | 66.1 | 47.3±22.2 |
|  | Change | 9.4 | 4.6 | 12.0 | 6.8 | 14.1 | 9.4±3.8 |
| CADL | Baseline | 42 | 57 | 34 | 40 | 51 | 51.4±20.5 |
|  | Week-3 | 49 | 67 | 39 | 48 | 59 | 61.8±22.9 |
|  | Change | 7 | 10 | 5 | 8 | 8 | 10.4±3.0 |
| UEFM | Baseline | 9 | 15 | 19 | 11 | 23 | 15.4±5.7 |
|  | Week-3 | 11 | 15 | 17 | 10 | 20 | 14.6±4.1 |
|  | Change | 2 | 0 | -2 | -1 | -3 | -0.8±1.9 |
| MAS-Elbow | Baseline | 1 | 2 | 1+ | 1 | 2 | 1(2),1+(1),2(2) |
|  | Week-3 | 0 | 1+ | 1+ | 0 | 1 | 0(2), 1+(2),1(1) |
|  | Change | 1 | 0.5 | 0 | 1 | 1 | 0(1),0.5(1),1(3) |
| MAS-Wrist | Baseline | 1 | 2 | 1+ | 2 | 1+ | 1(1),1+(2),2(2) |
|  | Week-3 | 1 | 2 | 1 | 0 | 1 | 0(1), 1(3), 2(1) |
|  | Change | 0 | 0 | 0.5 | 2 | 0.5 | 0(2),0.5(2),2(1) |
| MAS-Thumb | Baseline | 1 | 2 | 1 | 1+ | 1 | 1(3),1+(1),2(1) |
|  | Week-3 | 1 | 2 | 1 | 1 | 0 | 0(1),1(3),2(1) |
|  | Change | 0 | 0 | 0 | 0.5 | 1 | 0(3),0.5(1),1(1) |
| MAS-Fingers | Baseline | 1 | 1+ | 1 | 1+ | 2 | 1(2),1+(2),2(1) |
|  | Week-3 | 1 | 1+ | 0 | 1+ | 2 | 0(1),1(1),1+(2),2(1) |
|  | Change | 0 | 0 | 1 | 0 | 0 | 0(4),1(4) |

**Table 3.**
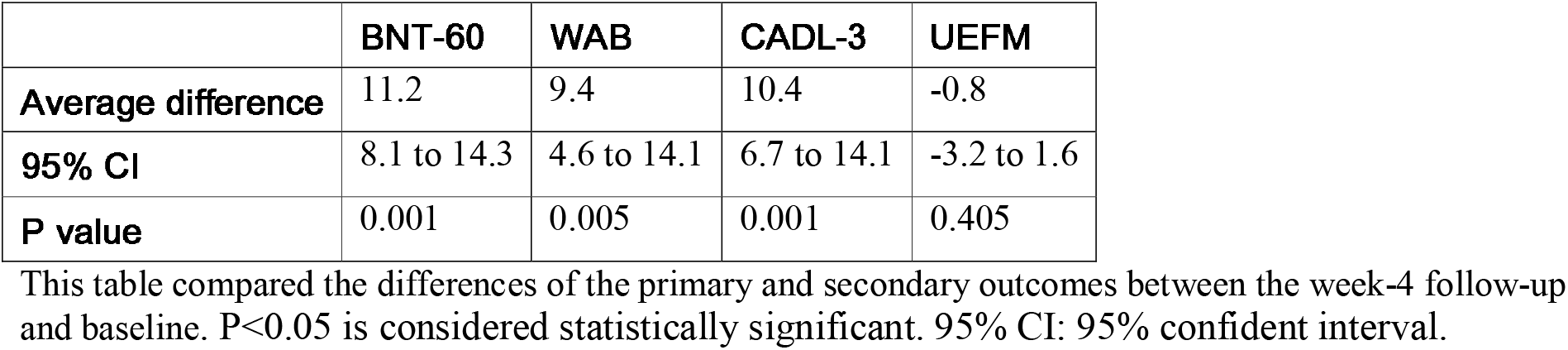
Statistics of the changes of the primary and secondary outcomes.

**Table 4.** Safety outcomes and side-effects.

| **Events** | **1** | **2** | **3** | **4** | **5** | **Total %** |
| --- | --- | --- | --- | --- | --- | --- |
| **Complications related to treatment**<br>**– no. of events** |  |  |  |  |  |  |
| Bleeding | N | N | N | N | N | 0 |
| Infection | N | N | N | N | N | 0 |
| Pain | Y | Y | N | Y | Y | 80% |
| Transient palsy of the phrenic nerve | N | N | N | N | N | 0 |
| **Changes in sensorimotor function**<br>**– no. of events** |  |  |  |  |  |  |
| Numbness | Y | Y | Y | Y | Y | 100% |
| Decrease in muscle strength-paralyzed side |  |  |  |  |  |  |
| Elbow | Y | Y | N | Y | Y | 80% |
| Wrist | Y | N | Y | N | N | 40% |
| Fingers | N | N | N | N | N | 0 |
| Tactile sensory threshold increases in thumb, index and middle fingers | Y | Y | Y | Y | Y | 100% |

Y refers to the presence of the incident in this patient, while N refers to the opposite.

## DISCUSSION

In this paper, we confirm the effectiveness of NC7 in enhancing language function by comparing the language improvement of five patients who received NC7 and iSLT in a prospective cohort study of NC7. For this result, our hypothesis included two possible mechanisms. The first one is the disruption of interhemispheric balance by NC7, altering efficacy in the right hemisphere. Research has shown that the left hemisphere of the brain is typically responsible for language processing in right-handed individuals [21]. However, the left hemisphere may become damaged for stroke, leading to aphasia. In some cases, the right hemisphere of the brain may be able to compensate [22, 23] for the loss of function by taking on some of the language processing tasks. It is thought that the enhanced function of the language network in the right hemisphere may be due to the increased activity and connectivity of the neurons in that hemisphere following the nerve transplant.

Another possible mechanism is that transection of the right C7 nerve root, especially near the intervertebral foreman which is close to the dorsal ganglion for delivering sensory input to the left hemisphere, will largely induce the functional plasticity of the left sensorimotor cortex. Since the sensorimotor cortex, especially the premotor cortex, is very close and even overlapped with the Broca’s area, which is an essential area for the language production, this large functional plasticity may interact with the left Broca’s area and induce benign plasticity which is beneficial to the language function.

Both of these two mechanisms could be a possible candidate after NC7, or were contributing simultaneously for the improvement of language function. To elucidate the underlying mechanism of language improvement after NC7, further exploration with functional MRI or electroencephalograph is needed. A preliminary exploration through event-related functional are performed in this cohort but not analysed in this article. Further research is needed to confirm both mechanisms to fully understand the mechanisms by which NC7 surgery improves language function in patients with aphasia.

## CONCLUSION

C7 neurotomy at the intervertebral foramen as a novel surgical approach, can improve the language function in adult patients with post-stroke aphasia. The underling mechanisms of the cortical reorganization still need further investigation.

## LIMITATION

The major limitation of this study is the study design as a preliminary observation. Randomized controlled clinical trial should be performed and more clinical evaluations as well as clinical neurophysiological studies should be added in further studies.

## Data Availability

All data produced in the present study are available upon reasonable request to the authors.

## REFERENCE

1. Stefaniak, J.D., A.D. Halai, and M.A. Lambon Ralph, The neural and neurocomputational bases of recovery from post-stroke aphasia. Nat Rev Neurol, 2020. 16(1): p. 43–55.

2. Fridriksson, J., et al., Anatomy of aphasia revisited. Brain, 2018. 141(3): p. 848–862.

3. Zhao, X., et al., Network-based Statistics Distinguish Anomic and Broca Aphasia. ArXiv, 2023.

4. Fridriksson, J. and A.E. Hillis, Current Approaches to the Treatment of Post-Stroke Aphasia. J Stroke, 2021. 23(2): p. 183–201.

5. Breitenstein, C., et al., Intensive speech and language therapy in patients with chronic aphasia after stroke: a randomised, open-label, blinded-endpoint, controlled trial in a health-care setting. Lancet, 2017. 389(10078): p. 1528–1538.

6. Sakamoto, R., et al., Intensive speech and language therapy after stroke. Lancet, 2017. 390(10091): p. 228.

7. Gholami, M., N. Pourbaghi, and S. Taghvatalab, Evaluation of rTMS in patients with poststroke aphasia: a systematic review and focused meta-analysis. Neurol Sci, 2022. 43(8): p. 4685–4694.

8. Biou, E., et al., Transcranial direct current stimulation in post-stroke aphasia rehabilitation: A systematic review. Ann Phys Rehabil Med, 2019. 62(2): p. 104–121.

9. Hope, T.M.H., et al., Right hemisphere structural adaptation and changing language skills years after left hemisphere stroke. Brain, 2017. 140(6): p. 1718–1728.

10. Zheng, M.X., et al., Trial of Contralateral Seventh Cervical Nerve Transfer for Spastic Arm Paralysis. N Engl J Med, 2018. 378(1): p. 22–34.

11. Feng, J., et al., Reconstruction of paralyzed arm function in patients with hemiplegia through contralateral seventh cervical nerve cross transfer: a multicenter study and real-world practice guidance. EClinicalMedicine, 2022. 43: p. 101258.

12. Peacock, W.J., L.J. Arens, and B. Berman, Cerebral palsy spasticity. Selective posterior rhizotomy. Pediatr Neurosci, 1987. 13(2): p. 61–6.

13. Albright, A.L., et al., Effects of continuous intrathecal baclofen infusion and selective posterior rhizotomy on upper extremity spasticity. Pediatr Neurosurg, 1995. 23(2): p. 82–5.

14. Cheung, R.W., M.C. Cheung, and A.S. Chan, Confrontation naming in Chinese patients with left, right or bilateral brain damage. J Int Neuropsychol Soc, 2004. 10(1): p. 46–53.

15. Chen, T.B., et al., Culture qualitatively but not quantitatively influences performance in the Boston naming test in a chinese-speaking population. Dement Geriatr Cogn Dis Extra, 2014. 4(1): p. 86–94.

16. Clark, H.M., et al., Western Aphasia Battery-Revised Profiles in Primary Progressive Aphasia and Primary Progressive Apraxia of Speech. Am J Speech Lang Pathol, 2020. 29(1s): p. 498–510.

17. Holland, A.L., D. Fromm, and L. Wozniak, CADL-3: Communication Activities of Daily Living.

18. Lin, Y.L., et al., Stratifying chronic stroke patients based on the influence of contralesional motor cortices: An inter-hemispheric inhibition study. Clin Neurophysiol, 2020. 131(10): p. 2516–2525.

19. Flanigan, M., et al., Spasticity and pain in adults with cerebral palsy. Dev Med Child Neurol, 2020. 62(3): p. 379–385.

20. Boardman, J.P., et al., Magnetic resonance image correlates of hemiparesis after neonatal and childhood middle cerebral artery stroke. Pediatrics, 2005. 115(2): p. 321–6.

21. Voets, N.L., et al., Distinct right frontal lobe activation in language processing following left hemisphere injury. Brain, 2006. 129(Pt 3): p. 754–66.

22. Balaev, V., A. Petrushevsky, and O. Martynova, Changes in Functional Connectivity of Default Mode Network with Auditory and Right Frontoparietal Networks in Poststroke Aphasia. Brain Connect, 2016. 6(9): p. 714–723.

23. Riès, S.K., N.F. Dronkers, and R.T. Knight, Choosing words: left hemisphere, right hemisphere, or both? Perspective on the lateralization of word retrieval. Ann N Y Acad Sci, 2016. 1369(1): p. 111–31.

